# Impact of COVID-19-lockdown and vulnerability factors on cognitive functioning and mental health in Italian population

**DOI:** 10.1101/2020.10.02.20205237

**Authors:** Eleonora Fiorenzato, Silvia Zabberoni, Alberto Costa, Giorgia Cona

**Affiliations:** Department of General Psychology, University of Padua, Padua, Italy; IRCCS Fondazione Santa Lucia, Rome, Italy; Niccolò Cusano University, Rome, Italy; Padova Neuroscience Center, University of Padua, Padua, Italy

**Author notes:** Corresponding author (GC).

## Abstract

The COVID-19 outbreak and its associated restrictive measures, such as lockdown, exposed most individuals to an unprecedented stressful situation, increasing worldwide the prevalence of mental health disorders.

Here, we aimed at exploring for the first time the effect of COVID-19-lockdown on subjective cognitive functioning as well as mental health, in terms of its prevalence and clinical severity. In addition, potential risk factors characterizing more vulnerable groups were investigated. A nationwide cross-sectional online-survey was conducted during the final phase of lockdown in Italy (after 7-10 weeks of home confinement), leading to a sample of 1215 participants.

We found lockdown can have a severe impact on subjective cognitive functioning, along with mental health disorders. Namely, under restrictions, cognitive complaints were mostly perceived in everyday tasks involving attention, temporal orientation and executive functions; while no changes in language abilities were reported. Conversely, a paradoxical effect was observed in memory domain, with people experiencing a reduction of memory failures compared to pre-lockdown times.

Further, higher severity and prevalence of depression, anxiety disorders and of other psychological issues (involving sleep, appetite, libido and hypochondria) were observed – with a prevalence of mild-to-severe depression and anxiety rising up to 32.30% and 35.72% respectively, under restrictions.

Being female, young (<45 years), repeatedly exposed to COVID-19-media, working from home or being unemployed were identified as relevant risk factors for experiencing cognitive worsening and mental health disorders, related to the COVID-19 lockdown. Being resident in high infection-prevalence areas was related to higher level of depression and health anxiety. As lockdown and quarantine measures could be reimposed, health officials have to carefully consider these more vulnerable groups in their decision-making process, to develop an effective global and long-term response to the mental health challenges of this pandemic as well as to implement psychological interventions and specific guidelines, particularly about COVID-19-media exposure.

## INTRODUCTION

Since the coronavirus disease 2019 (COVID-19) outbreak in December 2019, lockdown and mass quarantine measures have been adopted worldwide to limit the viral transmission and reduce the impact on healthcare systems. Italy was the first European country to apply a nationwide lockdown, which started on March 9 and ended in mid-May, requiring the population to home confinement and social isolation, with individuals being allowed to leave home only for limited and documented necessities (i.e., health reasons, shopping for essentials and work). Psychological and social consequences associated to this stressful and unique situation had been expected to be pervasive – affecting mental health under the pandemic restrictions, but also having long-term effects [1].

Evidence of quarantine from past viral outbreaks reported a massive impact on mental-health; albeit, those studies were mostly focused on medical staff and virus-affected patients with a quarantine no longer than 14 days [2].

In this regard, the exposure to COVID-19 related confinement seems to be an unprecedented condition, which cannot be compared with previous outbreaks as these differed in several aspects, above all for its worldwide magnitude. This global long-term home confinement of masses of people (to prevent COVID-19 from spreading), although with access to digital tools to ensure the maintenance of education, work, social interaction and communication, led to a global atmosphere of depression, anxiety and stress associated to home-confinement, social isolation, disrupted travel, media information overload and panic for essential items purchase [3–5].

The psychological response during the COVID-19 lockdown has been studied since the initial outbreak [5–9]; however, data are conflicting particularly about the vulnerability factors that might predict mental health outcomes in the general population. Although leading to controversial findings, the most relevant vulnerability factors, identified by previous studies, included: gender [7,10], age [5,7,11,12], occupational status [7,8], exposure to social media and COVID-related information [11,13] as well as territory of residency [14]. Of note, some of this previous research used brief screening scales (e.g., 2-items) to assess depression and anxiety that did not allow to capture symptoms severity [7], which is an important aspect to compare results across studies [15]. Finally, despite the effect of COVID-19 lockdown on sleep disorders having been extensively studied [3], only a little evidence on other psychological issues (e.g., eating, sexual disorders and anxiety for health) has been reported. Notably, the impact of COVID-19 home confinement on subjective cognitive functioning remained an unexplored research topic; however, answering this research question seems particularly relevant given that subjective cognitive complaints have been recognized to significantly predict everyday-task functioning [16,17].

Considering the strong impact on mental health outcomes associated to this unprecedent stressful condition, in the present work for the first, time we want to address possible consequences of lockdown on subjective cognition.

Given the above, our study aims at exploring the psychological impact of COVID-19-restrictive measures on subjective cognitive functioning as well as mental health in the final phase of lockdown in Italy (i.e., after 7-10 weeks of lockdown), and at identifying risk factors that characterize potential vulnerable groups among the general population. To achieve this, we administered a nationwide cross-sectional online-survey to reach a large cohort and ensure an adequate representation of the Italian general population.

In particular, we first aimed to explore the presence of subjective cognitive complaints perceived under restrictions, as compared to normal times (before the lockdown). Focusing on possible changes in attention, executive functions, memory, language abilities and temporal orientation.

Further, our second aim was to determine the prevalence and severity of mental health disorders, as well as of other psychological issues (i.e., sleep, appetite, libido and anxiety for health) associated to lockdown restrictions as compared to before. Our main hypothesis was that COVID-19 lockdown might have a detrimental effect on subjective cognitive functioning, converging with previous evidence observed about mental health changes.

In addition, another purpose was to identify the vulnerability factors that might influence subjective cognition as well as mental health outcomes in this unique context. Indeed, our final aim was to identify potentially vulnerable groups or possible risk factors in order to pave the way to implement specific interventions and reduce the burden of psychological issues related to COVID-19 restrictions.

We believe our results can assist government decision-making and healthcare professionals in developing a global and long-term response to cognitive and mental health challenges of this pandemic.

## METHODS

### Study design and participants

An anonymous online survey was shared through various platforms and mainstream social media for a limited time window, from April 29 to May 17, 2020. This timeframe was chosen to assess participants’ response during the final phase of the COVID-19 pandemic lockdown, following the Italian government declaration of lockdown (Decree of March 9, 2020) and the national reopening announcement (May 18, 2020).

To obtain a representative countrywide sample of the Italian population, which has been differently affected by the pandemic, a snowball sampling method was used. Participants, in addition to their own contribution, were encouraged to share and invite new respondents among their contacts. Of note, we emphasized the involvement of the elderly and people with a poor access to internet, encouraging participants to help them filling out the survey. Participation was voluntary and without compensation.

A brief introduction informed the participants about our study’s aims and their informed consent was requested before starting the investigation. The survey took approximately 20 minutes and was anonymous, assuring data confidentiality.

Responses were considered eligible if participants: i) completed the entire survey, ii) were aged over 18 years old and iii) were living in Italy during the pandemic.

Among a total of 1559 responses via Qualtrics platform, 1242 were classified as eligible according to our inclusion criteria and from this sample we excluded 27 participants, who reported chronic neurological conditions (i.e., neurodegenerative disease, traumatic brain injury) or a history of mental disorders. The final sample consisted of 1215 participants.

The study was conducted in accordance with the Helsinki Declaration and approved by the ethical committee of the School of Psychology (University of Padua), and Fondazione S. Lucia, Rome.

### Survey structure and outcome measures

The survey self-report questionnaire was composed of three sections. The first section was set up to collect sociodemographic features and COVID-19 related information. In the second and third sections, we presented two identical self-reported questionnaires (organized in four subsections), wherein participants were asked to fill out the questionnaires thinking about their condition in normal times (pre-lockdown) vs. during the lockdown. Specifically, the second section comprised questions about a typical week before COVID-19 outbreak (such as during the first week of February — from Monday, 3 to Sunday, February 9, 2020). While in the third section, participants were asked to reply by thinking of their condition during the last week of the COVID-19 lockdown (such as during the last week of April — from Monday April 27 to Sunday May 3, 2020).

Second and third sections were both organized in four subsections, investigating: i) daily life habits (e.g., average hours spent at home, at workplace or doing sport, number of people living together, drug and psychotropic drug use), ii) subjective memory abilities, iii) subjective global cognitive functioning, and iv) presence of anxiety, depression and other psychological issues (such as sleep disorders, changes in appetite, libido and hypochondria).

#### Sociodemographic, COVID-19 related information and daily life habits

A dedicated questionnaire was set up to collect sociodemographic variables of interest, while the COVID-19 section aimed to collect information on lockdown conditions (working status, living condition, need of psychological consultation, number of time outside home for a walk or shopping.) and about COVID-19 (contraction of COVID-19 infection and related symptoms, fear, contact with confirmed cases, media consumption about COVID-19).

#### Memory functioning

The Prospective and Retrospective Memory Questionnaire (PRMQ) was used to assess memory slips that everyone can make in daily life [18]. PRMQ is a 16-item set, where participants are asked to report on a 5-point scale how frequently they experienced some memory mistakes, ranging from ‘very often’ to ‘never’. Higher PRMQ total scores are suggestive of more frequent self-reported memory difficulties (max-min total score: 80-16). Of note, we slightly modified only Item 12, since it was an unsuitable activity during the lockdown (‘do you fail to mention or give something to a visitor’). The item was rephrased as follow ‘do you fail to mention or say something to someone that you had contacted’.

#### Perceived cognitive functioning

An ad-hoc 10-item questionnaire was created to assess the subjective global cognitive functioning in performing everyday tasks, identified as feasible activities in a home confinement condition. Items were derived from standardized tools used in clinical practice to assess subjective cognitive complaints: namely, the Perceived Deficits Questionnaire [19] and the Cognitive Change Questionnaire [20]. Our 10-item questionnaire aims to assess perceived cognitive problems (e.g. “I have trouble concentrating. For instance, while reading, watching a TV program, working”, “I have trouble doing multiple things at once”) by capturing subtle difficulties in performing activities of daily living, which are related to possible cognitive changes.

The investigated everyday tasks requires *attention and concentration* abilities (three items referring to difficulties in concentrating such as attention while watching a TV program, talking or looking for an object at home), *executive functions* (three items referring to difficulties in multitasking, problem-solving and decision-making), *temporal orientation* (two items referring to the ability of tracking the time such as remembering the day of the week/month, an important date, as a birthday) and *language abilities* (two items referring to the ability of finding the proper word and express themselves) (for further details see S1 Appendix). Participants were asked on a 5-point scale how frequently they experienced each of the difficulties ranging from very often to never. Total score ranges from 10 to 50, with higher scores representing greater perceived cognitive impairment. A total score as well as subscores for each cognitive domain were computed.

#### Depression and anxiety

The Hospital Anxiety and Depression Scale (HADS) was used to assess presence of anxious and depressive symptoms [21]. HADS is a brief 14-item self-rating scale widely used instrument to detect states of anxiety and depression in general practice with excellent psychometric properties [22] and validated in Italian. HADS is composed of two scales to assess anxiety and depression (HADS-A and HADS-D, respectively), yielding two separate measures of distinct emotional disturbances [23]. Higher total score indicates higher severity in terms of anxiety and depressive symptoms. To identify presence of clinically significant anxiety or depression disturbances, a cutoff score of 8 was adopted (ranges were defined as follow: 8-10 mild, 11-14 moderate, above 15 severe symptoms) [22].

#### Sleep, appetite, libido and hypochondria changes

To measure changes in appetite, sleep and interest in sex, we included three Beck Depression Inventory (BDI-II) items (16, 18 and 21, respectively) [24], which we modified in order to capture changes bidirectionally as behavioral increment or decrement (e.g., increase vs. loss of appetite). Participants were asked to report presence of mild, moderate or significant changes in these dimensions on a 4-point scale, where higher score were suggestive of greater changes. Finally, an item about hypochondria was included. This 4-point scale item of the Hamilton Depression Rating Scale [25] assessed presence of anxiety for health, where a higher score indicates more severe symptoms.

### Statistical analysis

Descriptive analyses were performed for all outcome measures, the Kolmogorov-Smirnov test was used to test normality of continuous variables, while the Levene test for the homogeneity of variance was employed. Categorical variables were analyzed through Chi-square test or McNemar’s test, when comparing repeated measures. Correlations analyses were computed to explore the relationship between variables of interest (e.g., mood/anxiety changes).

Based on previous evidence [7,11,14] we identified five factors that were particularly relevant to characterize potential vulnerable groups: gender, age, occupational status, COVID-19-media exposure and territory of residency (North vs. South of Italy – due to the different incidence within the country, namely with the northern regions being the most affected by the infection).

Hence, first, we verified the effect of the five vulnerability factors on cognitive and mental health changes (related to the lockdown), by means of multi-factorial ANOVA. Score change was defined as score reported under restrictions subtracted by the score before the lockdown. Subsequently, those factors resulting significant in multi-factorial models were better explored through separate repeated-measure ANOVAs (RM-ANOVA) and included as between-subject factors — whereas, pre- and during-lockdown cognitive and mental health outcomes were set as within-subject factors. Our main goal was to explore the interaction of Lockdown x vulnerability factors, and further through post-hoc analyses to identify the most vulnerable subgroups.

In addition, age and/or education were entered as covariates in RM-ANOVA, if groups were not matched for these variables. Regarding the analyses on cognitive measures, we checked the possible confounding effect of depressive mood and anxiety changes on subjective cognitive functioning by entering those variables as covariates in RM-ANOVAs. Following this check, if results did not change, the variables were then removed from the covariates to not lose statistical power.

Statistical significance threshold was set at p < .05 and post-hoc were conducted applying the Holm-Bonferroni correction for multiple comparisons. When necessary, planned comparisons were run to better explore the predictive interactions effects yielded by RM-ANOVA. Effect sizes were estimated using partial eta squared (ηp^²^). We performed statistical analyses using SPSS Statistic, release version 24.0 (Chicago, IL, USA).

## RESULTS

### Sociodemographic and COVID-19 related features

Total sample (n=1215) sociodemographic characteristics as well as COVID-19 related information are shown in Table 1.

**Table 1.**
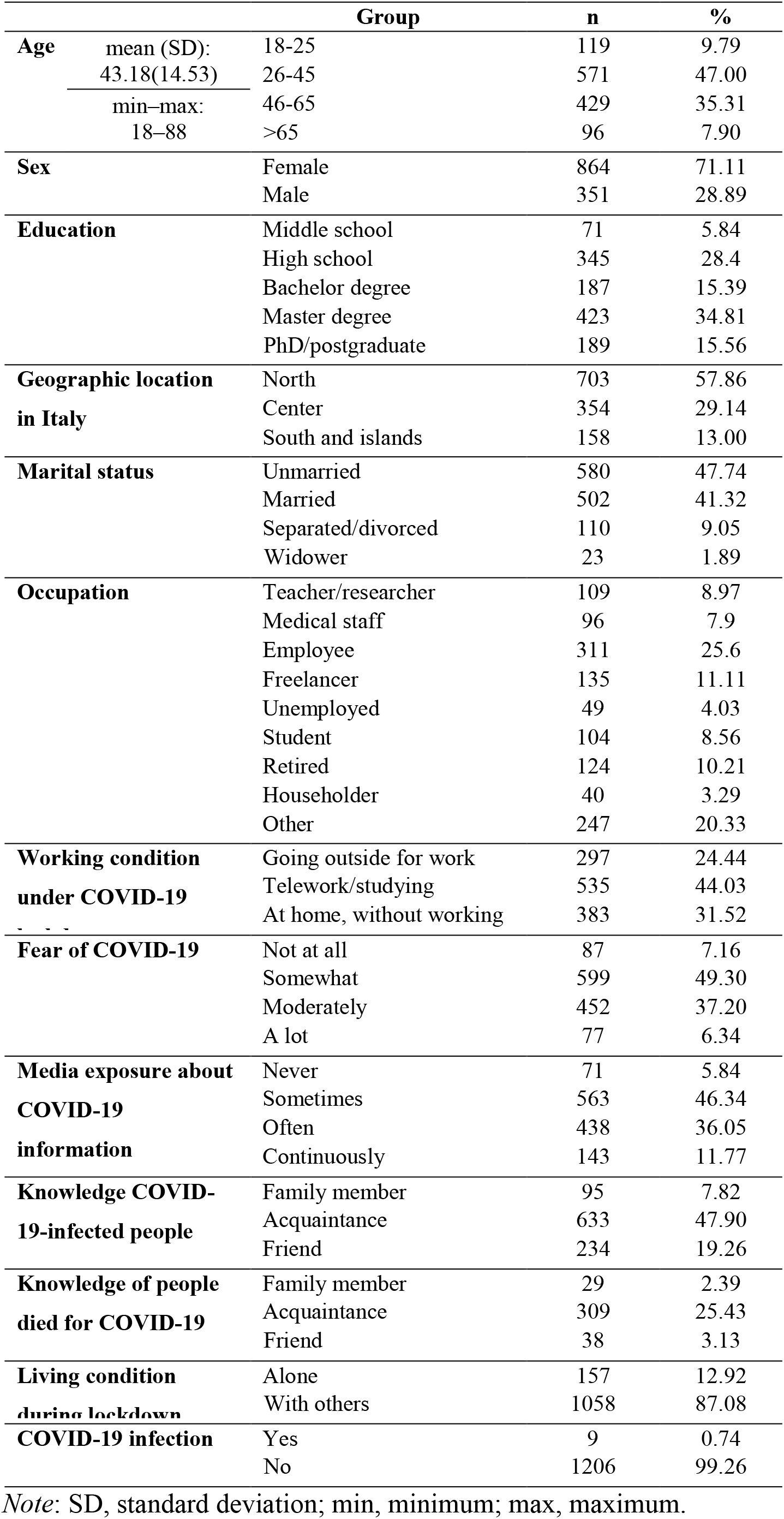
Total sample sociodemographic characteristics and COVID-19-related information

### Lockdown effect on subjective cognitive functioning and mental health

As shown in Table 2, RM-ANOVA results revealed that under lockdown-restrictions, our sample perceived a worsening on global cognitive functioning as compared to normal times (pre-lockdown) (p < .001), but an improvement on memory abilities (p < .001). In particular, PRMQ revealed a reduction of everyday-life memory slips, in both prospective and retrospective memory components. Of note, these findings on subjective cognitive functioning were confirmed and resulted statistically significant also when mood and anxiety changes were entered as covariates in RM-ANOVA.

**Table 2.** Subjective cognitive functioning and mental health before vs. during COVID-19 lockdown, in the total sample (n = 1215).

| | Pre-lockdown | During-lockdown | F/$\chi^2$ | p | $\eta_p^2$ |
| --- | --- | --- | --- | --- | --- |
| ***Cognitive measures*** |  |  |  |  |  |
| PRMQ (total score) | 32.61 (8.90) | 30.98 (10.24) | 55.25 | < .001 | .04 |
| Prospective memory | 17.45 (4.92) | 16.51 (5.71) | 53.12 | < .001 | .04 |
| Retrospective memory | 15.16 (4.42) | 14.48 (4.98) | 42.43 | < .001 | .03 |
| Subjective cognitive | 19.99 (5.79) | 21.13 (7.45) | 45.88 | < .001 | .04 |
| ***Mental health measures*** |  |  |  |  |  |
| HADS-D (mean, SD) | 3.83 (3.33) | 5.81 (3.97) | 338.36 | < .001 | .22 |
| **cutoff $\geq 8$ (n)** | **15.39% (n=187)** | **32.30% (n=392)** | 125.73 | < .001 |  |
| mild (8–10) | 11.53% | 19.03% |  |  |  |
| moderate (11–14) | 4.03% | 10.89% |  |  |  |
| severe ($\geq 15$) | 0.32% | 2.82% | | | |
| HADS-A (mean, SD) | 5.21 (3.23) | 6.51 (4.03) | 115.08 | < .001 | .11 |
| **cutoff $\geq 8$ (n)** | **21.40% (n=260)** | **35.72% (n=434)** | 86.00 | < .001 |  |
| mild (8–10) | 15.24% | 19.19% |  |  |  |
| moderate (11–14) | 5.48% | 12.10% |  |  |  |
| severe ($\geq 15$) | 1.21% | 4.76% | | | |
| Sleep (mean, SD) | 0.50 (0.83) | 0.94 (0.9) | 213.517 | < .001 | .15 |
| % reporting a change | 33.25% (n=404) | 63.95% (n=777) | 272.95 | < .001 |  |
| increase vs. decrease | 102/302 | 360/417 |  |  |  |
| Appetite (mean, SD) | 0.31 (0.67) | 0.67 (0.85) | 212.886 | < .001 | .15 |
| % reporting a change | 22.47% (n= 273) | 48.15% (n=585) | 240.60 | < .001 |  |
| increase vs. decrease | 190/83 | 387/198 |  |  |  |
| Interest in Sex (mean, SD) | 0.39 (0.76) | 0.57 (0.84) | 68.873 | < .001 | .05 |
| % reporting a change | 26.01% (n= 316) | 38.70% (n= 471) | 80.94 | < .001 |  |
| increase vs. decrease | 72/244 | 169/302 |  |  |  |
| Hypochondria (mean, SD) | 0.38 (0.71) | 0.49 (0.78) | 35.136 | < .001 | .03 |
| % reporting a change | 26.67% (n= 324) | 35.14% (n= 427) | 47.95 | < .001 |  |

*Note:* SD, standard deviation; PRMQ, Prospective and Retrospective Memory Questionnaire; HADS-D, Hospital Anxiety and Depression Scale for depression; HADS-A, HADS for anxiety; $\eta_p^2$ , partial eta squared. Continuous variables were compared by means of repeated-measure analysis of variance, while categorical variables through McNemar's test. Values in bold type indicate the overall prevalence according to HADS cut-off $\geq 8$ , followed by the specific prevalence according to severity (ranges were defined as: 8–10 mild, 11–14 moderate, above 15 severe) [22]. The referred changes in sleeping pattern, appetite, libido and hypochondria were defined as having a score $\geq 1$ . Further, sleep, appetite, libido changes were characterized based on the behavior increase vs. decreased (e.g., increase vs. loss of appetite). Higher total score indicates higher severity in terms of cognitive and mental health symptoms.

Looking at mental health changes (pre-vs. during-lockdown), we found a significant increase of HADS-D and HADS-A scores (as shown in Table 2) suggesting an overall increase in depressive and anxiety disorders related to lockdown. In particular, under restrictions, the prevalence of people reporting clinically relevant (cut-off score **≥** 8) depressive symptoms raised up to 32.30% as compared to normal times (15.39%) (χ^2^_1_= 125.73, p < .001; Table 2), of whom 13.71% and 4.35% showing moderate/severe levels of depression, respectively (χ^2^_1_= 64.76, p < .001; Table 2)

Similarly, clinically relevant (cut-off score ***≥*** 8) anxiety disorders were present in 35.72% of our sample compared to the 21.40% under normal times (χ^2^_1_= 86.00, p < .001; Table 2), of whom 16.86% and 6.69% showed moderate/severe levels of anxiety, respectively (χ^2^_1_= 60.46, p < .001; Table 2).

As reported in Table 2, concerning other psychological issues, we found that restrictions led to an increase in problems related to sleep, appetite, libido and hypochondria as compared with pre-lockdown (p < .001). In particular, changes in sleeping pattern were reported by the 63.95% (n=777), of whom about 54% experiencing insomnia and the remaining increased sleepiness. For appetite score, the 48.15% (n=585) reported changes, mostly characterized by an increased appetite (∼66%, n= 387). Regarding changes in libido, these were reported by the 38.70% (n= 471), mostly characterized by a reduced interest in sex (∼64%, n=302). Finally, during the lockdown, the 35.14% (n= 427) showed an increased anxiety for health as compared to the 26.67% before the lockdown (p < .001).

Furthermore, as shown in S1 Table, according to multifactorial models results, we identified five vulnerability factors that were significant in explaining the lockdown effect on cognitive functioning and mental health, namely: gender, age, working condition during lockdown, geographical areas (North vs. Center-South of Italy), and media consumption about COVID-19 (i.e., media usage frequency). Thus, to better highlight the interaction as well as the individual profiles that are mostly affected by lockdown effect, RM-ANOVAs were run and results are reported as follows.

### Gender

RM-ANOVA results showed a significant Lockdown × Gender interaction on cognitive functioning (F_1,1212_ = 17.01, p < .001, η_p_^²^ = .014; Fig 1a), revealing women perceived a more pronounced worsening in cognitive functioning as compared to men. In line with this finding, a significant interaction was observed in the sub-items analysis (Fig 1b). As such, women experienced more difficulties in everyday tasks involving attention and concentration abilities (F_1,1212_ = 11.14, p < .001, η_p_^²^ = .009), executive functions (F_1,1212_ = 13.28, p < .001, η_p_^²^ = .011) and temporal orientation (F_1,1212_ = 19.32, p < .001, η_p_^²^ = .016); while no changes were perceived in language abilities (F_1,1212_ = 19.32, p < .001, η_p_^²^ = .016) during the lockdown. Of note, the interaction was confirmed also when mood and anxiety changes were entered as covariates.

**Fig 1.**
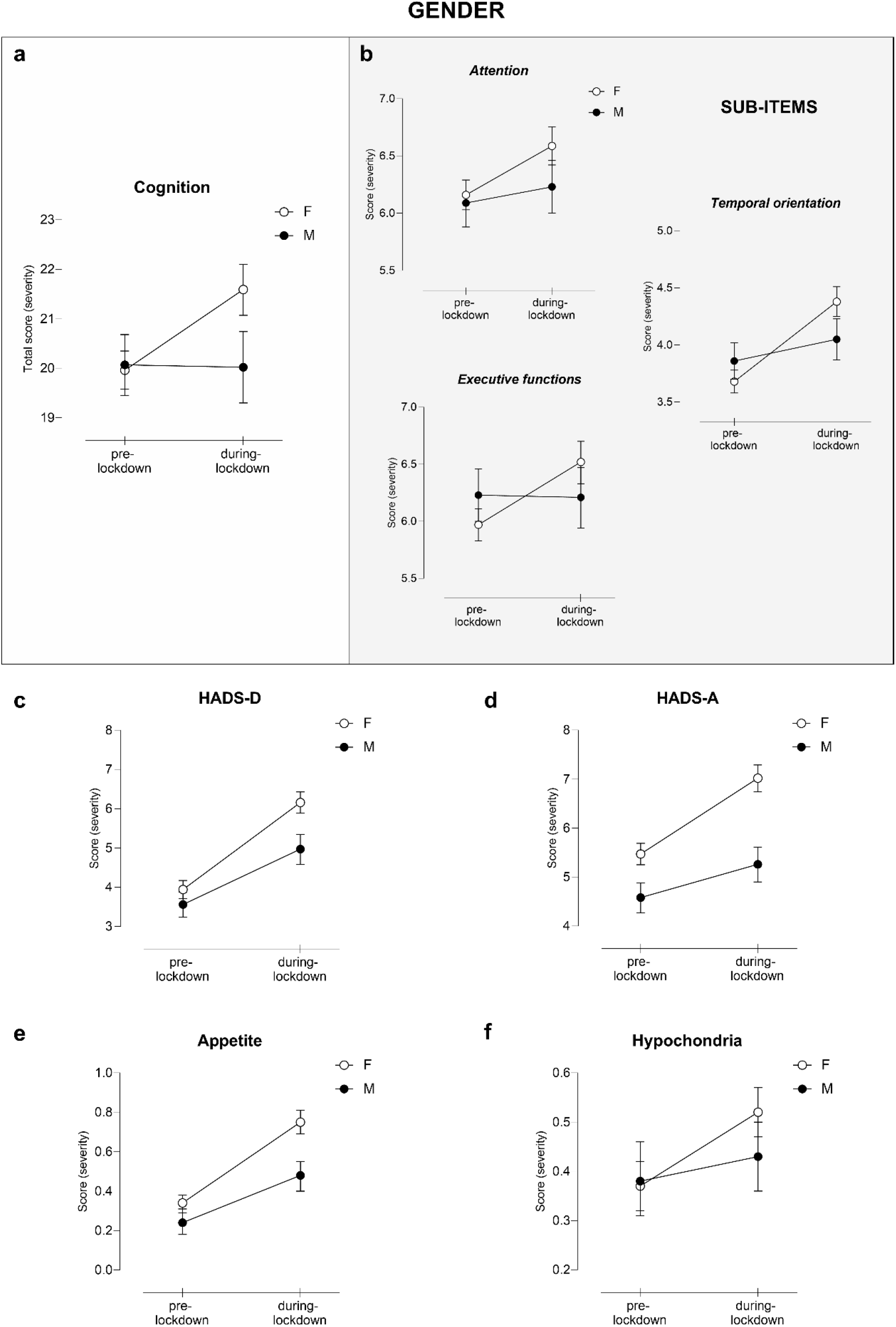
COVID-19-lockdown effect according to Gender. Changes in a) subjective cognitive functioning total score and b) its subitems, c) HADS-D, d) HADS-A, e) appetite and f) hypochondria as a function of Lockdown and Gender. Only significant interactions Lockdown x Gender are displayed, following RM-ANOVA analysis, entering age as covariate. Higher total score indicates higher severity in terms of cognitive and mental health symptoms. Error bars represent confidence intervals. HADS-D, Hospital Anxiety and Depression Scale for depression; HADS-A, HADS for anxiety.

Regarding mental health, we found a statistically significant Lockdown × Gender interaction in all the investigated outcomes. Lockdown induced a more significant increase in anxiety and depressive symptoms in women than men, as measured by HADS-D and HADS-A respectively (F_1,1212_ = 10.22, p < .001, η_p_^²^ = .008; F_1,1212_ = 13.43, p < .001, η_p_^²^ = .011) (Fig 1c and d). As shown in Fig 2a, under restrictions, the prevalence of anxiety disorders increases up to 40.51% in women and 23.93% in men (χ^2^_1_= 29.88, p < .001; log odds ratio .772 [CI = .48-1.05]), albeit in the context of a gender difference already present at the pre-lockdown assessment (χ^2^_1_= 9.63, p < .002). This result suggests that the probability of experiencing depressive disorders was 2.2-fold greater in women than in men during lockdown. Likewise, we found an interaction between lockdown and gender in depressive disorders, which increased up to 35.19% in women; whereas, to 25.07% in men (χ^2^_1_= 11.68, p < .001; log odds ratio .484 [CI = .21-.76]; Fig 2b); in the context of no differences at baseline (p = .159), suggesting that the probability of experiencing depression during the lockdown was 1.6-fold greater in women than in men.

**Fig 2.**
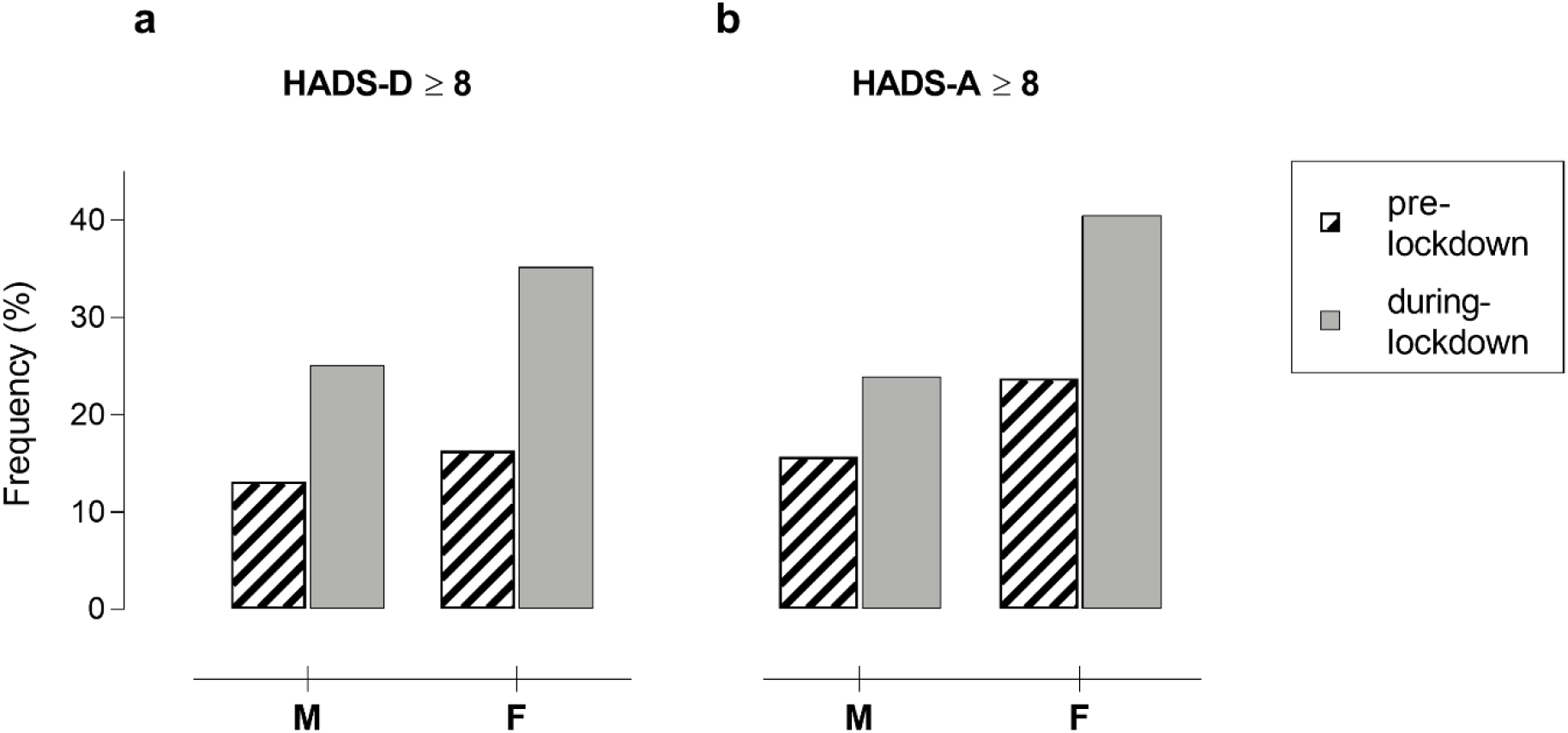
Prevalence of mild depression and anxiety according to Gender, pre- and during-lockdown. Mild symptoms assessed with a cut-off ≥8 of a) HADS-D and b) HADS-A scales. HADS-D, Hospital Anxiety and Depression Scale for depression; HADS-A, HADS for anxiety.

Furthermore, looking at the other psychological issues related to the lockdown, we found that women experienced a greater change in the appetite as compared to men (F_1,1212_ = 8.74, p = .003, η_p_^²^ = .007; Fig 1e), showing increased eating behaviors, and displayed a more pronounced increment in hypochondria and anxiety for health compared to men (F_1,1212_ = 4.50, p = .034, η_p_^²^ = .004; Fig 1f), whereas changes in sleeping patterns and libido were equally affected in both genders.

### Age

Looking at the changes on subjective cognitive functioning during lockdown, we found a Lockdown × Age interaction (F_1,1210_ = 13.97, p < .001, η_p_^²^ = .033; Fig 3a). Post-hoc comparisons revealed that the self-reported worsening of cognitive functioning was statistically significant only in the younger generations (18-25 and 26-45 years) (19.98 vs. 23.66; p < .001 and 20.32 vs. 21.89; p < .001, respectively). Sub-items analyses confirmed this result (Fig 3b), showing the perceived decline was driven particularly by daily life activities involving attention and concentration (F_1,1210_ = 7.88, p < .001, η_p_^²^ = .019; Fig 3b), executive functions (F_1,1210_ = 6.46, p < .001, η_p_^²^ = .016; Fig 3b) and temporal orientation (F_1,1210_ = 24.01, p < .001, η_p_^²^ = .056; Fig 3b), but not language abilities (F_1,1210_ = 3.98, p < .008, η_p_^²^ = .010). Of note, no interaction was found in the measure related to memory. About mental health, we found a statistically significant Lockdown × Age interaction in HADS-D (F_1,1211_ = 2.93, p < .033, η_p_^²^ = .007; Fig 3c), indicating the lockdown induced a significant increase in depressive symptoms. Since post-hoc results showed mood changes were significant in all age groups (p < .001), to better investigate this interaction, planned comparisons were conducted. Here, we found mood worsening was greater in the 26-45 age group than in the 46-65 age group (p = .022).

**Fig 3.**
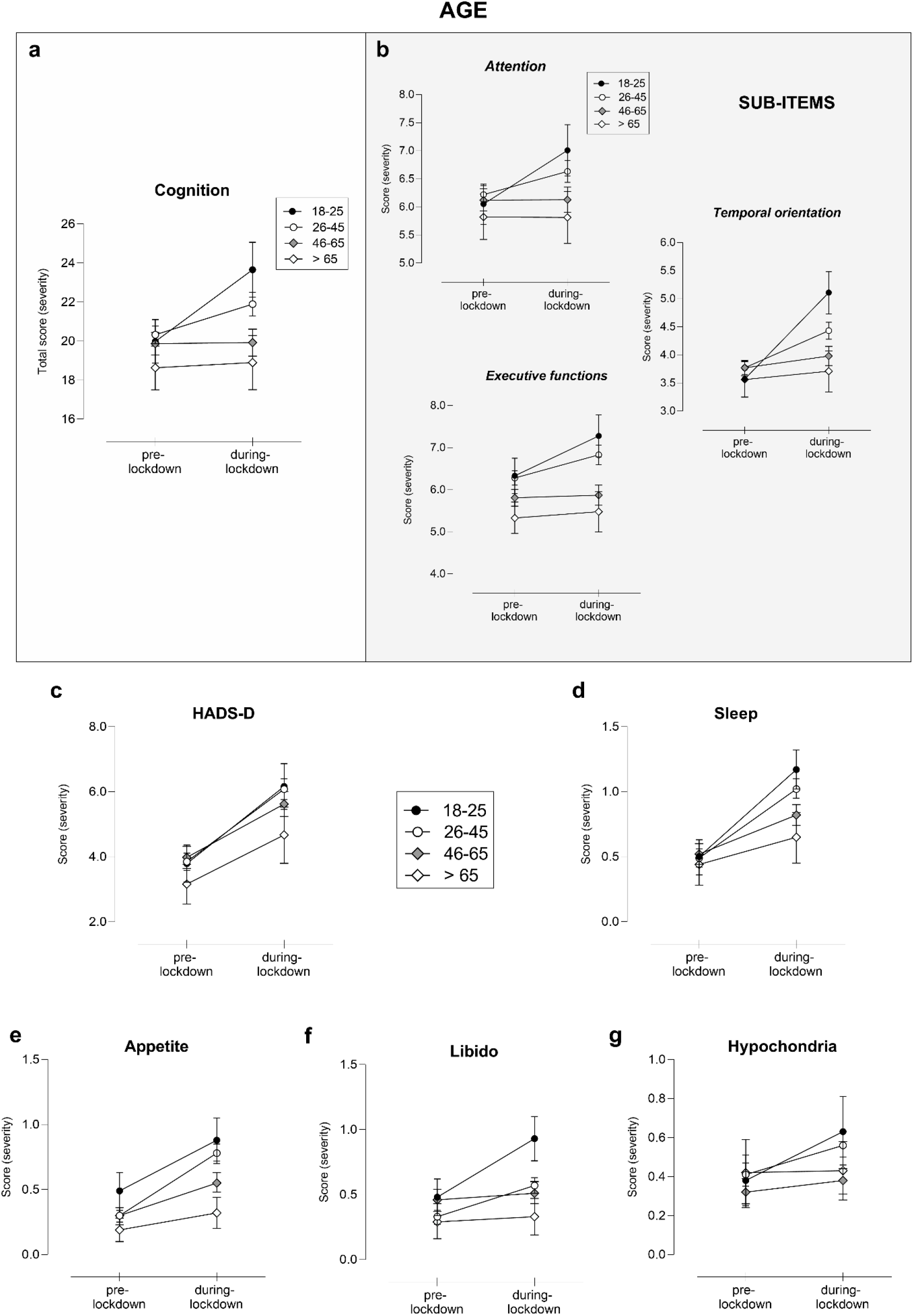
COVID-19-lockdown effect according to Age. Changes in a) subjective cognitive functioning total score and b) its subitems, c) HADS-D, d) sleep, e) appetite, f) libido and g) hypochondria as a function of Lockdown and Gender. Only significant interactions Lockdown x Age are displayed, following RM-ANOVA analysis, entering education as covariate. Higher total score indicates higher severity in terms of cognitive and mental health symptoms. Error bars represent confidence intervals. HADS-D, Hospital Anxiety and Depression Scale for depression.

Likewise, we found a Lockdown × Age interaction in sleep (F_1,1211_ = 7.72, p < .001, ηp^²^ = .019; Fig 3e), appetite (F_1,1211_ = 7.43, p < .001, η_p_^²^ = .018; Fig 3d), libido (F_1,1211_ = 11.55, p < .001, η_p_^²^ = .028; Fig 3f) and hypochondria (F_1,1211_ = 4.11, p = .007, η_p_^²^ = .010; Fig 3g). Post-hoc comparisons revealed greater changes in appetite (characterized by an increment) in the group aged 26-45 years as compared to the older generations (>45 years) (p < .001). Changes in sleep, libido and hypochondria were significantly greater in the younger (<45 years) as compared to the older generations (>45 years) (p < .010, p < .001 and p < .002, respectively). Interestingly, sleep changes in the younger population were both in terms of poor sleep as well as an increase sleepiness. Concerning changes in libido, we found that the youngest individuals (18-25 years) experienced an increased interest in sex, whereas the 26-45 group a reduction under the lockdown.

### Working condition

A statistically significant Lockdown × Working condition interaction on global cognition (F_1,1210_ = 3.13, p = .040, η_p_^²^ = .005; Fig 4a) was found. Post-hoc analysis following RM-ANOVA revealed no-working and teleworking groups perceived greater cognitive worsening during the lockdown, as compared to times pre-lockdown (21.56 vs. 20.38; p < .001 and 21.46 vs. 19.97; p < .001, respectively). Aligned with this result, a significant interaction was also observed in the sub-items analysis (Fig 4b), particularly in the attention and concentration domain (F_1,1210_ = 3.52, p = .030, η_p_^²^ = .006) and temporal orientation (F_1,1210_ = 5.38, p = .005, η_p_^²^ = .009).

**Fig 4.**
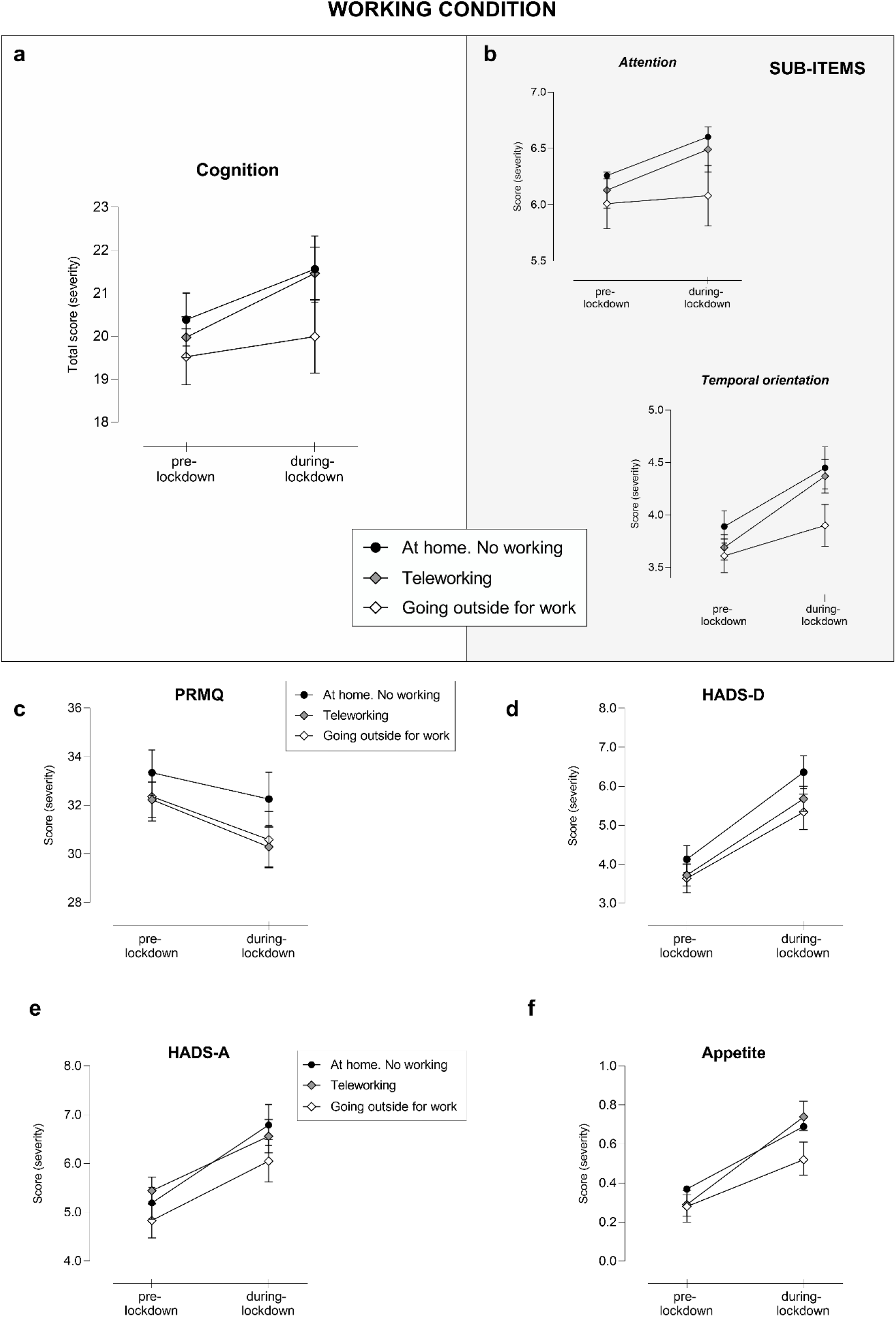
COVID-19-lockdown effect according to the Working condition. Changes in a) subjective cognitive functioning total score and b) its subitems, c) PRMQ, d) HADS-D, e) HADS-A and f) appetite as a function of Lockdown and Working condition. Only significant interactions Lockdown x Working condition are displayed, following RM-ANOVA analysis, entering age and education as covariates. Higher total score indicates higher severity in terms of cognitive and mental health symptoms. Error bars represent confidence intervals. PRMQ, Prospective and Retrospective Memory Questionnaire; HADS-D, Hospital Anxiety and Depression Scale for depression; HADS-A, HADS for anxiety.

Regarding memory abilities, we found a statistically significant interaction Lockdown × Working condition effect (F_1,1210_ = 3.02, p = .050, η_p_^²^ = .005; Fig 4c). More specifically, post-hoc analysis highlighted a perceived improvement in memory during the lockdown as compared to normal times in the two working groups (teleworking and outside home) (p < .001), while no differences were perceived by the no-working group (p = .321). Noteworthy, perceived improvement in memory domain was associated with a positive change of depressive mood (r = .34, p < .001) and anxiety disorders (r = .38, p < .001) (S1 Fig).

The RM-ANOVA on mental health outcomes showed a statistically significant Lockdown × Working condition interaction in HADS-D (F_1,1211_ = 3.35, p = .040, η_p_^²^ = .006; Fig 4d) and HADS-A (F_1,1211_ = 3.99, p = .020, η_p_^²^ = .007; Fig 4e) scales, wherein a significant different impact of lockdown on depressive and anxiety symptoms as a function of the Working condition was observed. As revealed by the post-hoc analysis, however, anxiety and mood changed in all working groups (p < .001). Therefore, to better explore this interaction, planned comparisons were computed. These analyses revealed the worsening in depressive symptoms was particularly pronounced in the no-working group, as compared to the two working groups (teleworking and outside home) (p = .030; p = .020, respectively); similarly, this pattern was observed for anxiety symptoms (p = .005; p = .050, respectively). Looking at the changes in the other psychological and habit dimensions, a Lockdown × Working condition interaction in appetite scores (F_1,1211_ = 6.43, p = .002, η_p_^²^ = .011; Fig 4f) was observed. Since post-hoc analysis was unable to capture this difference (the change was significant in all groups), planned comparisons were conducted – revealing the change was greater (in terms of increased appetite) in the teleworking group as compared to the no-working (p = .044) and working outside home (p = .002) groups. This result however has to be considered with caution.

### Italian geographical areas

RM-ANOVA results showed a statistically significant Lockdown × Geographical areas interaction on mental health outcomes, in particular on depressive symptoms as measured by HADS-D (F_1,1212_ = 8.48, p = .004, η_p_^²^ = .007; Fig 5a). We found that residents in Northern Italy perceived an increase in depressive disorders as compared to people living in Southern Italy during the lockdown. Likewise, when looking at changes in other psychological issues, an interaction was found in hypochondria associated with the lockdown (F_1,1212_ = 6.41, p = .010, η_p_^²^ = .005; Fig 5b), where again residents in Northern Italy reported greater health-related fears than the other group.

**Fig 5.**
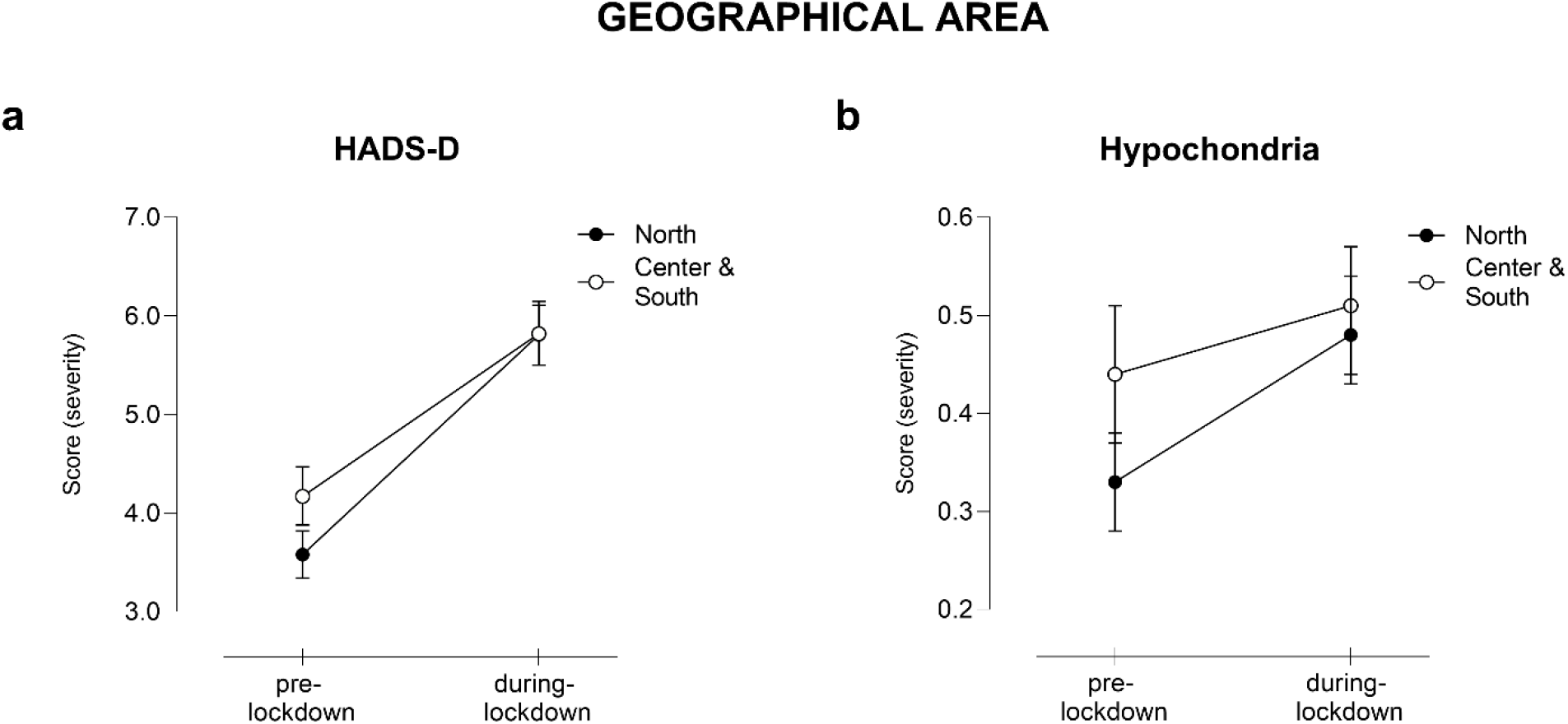
COVID-19-lockdown effect according to the Italian Geographical area. Changes in a) HADS-D and f) hypochondria as a function of Lockdown and Geographical area. Only significant interactions Lockdown x Geographical area are displayed, following RM-ANOVA analysis, entering age as covariate. Higher total score indicates higher severity in terms of mental health symptoms. Error bars represent confidence intervals. HADS-D, Hospital Anxiety and Depression Scale for depression.

### Media exposure about COVID-19

By categorizing our sample based on the frequency of media consumption about COVID-19 (i.e., media usage frequency) during the lockdown, we found a Lockdown × Media exposure interaction in several cognitive and mental health outcome measures (Fig 6). Namely, RM-ANOVA results showed a statistically significant interaction of Lockdown × Media exposure (F_1,1210_ = 3.25, p < .021, η_p_^²^ = .008; Fig 6a) on subjective cognitive functioning. Post-hoc comparisons revealed this cognitive worsening was mostly perceived by frequent media seekers (often/ continuously) (p < .001) rather than occasional media seekers (never/sometimes) (p > .050). Sub-items analyses confirmed this result (Fig 6b), revealing the perceived decline was driven particularly by attention and concentration (F_1,1210_ = 2.91, p = .030, η_p_^²^ = .007; Fig 6b) and temporal orientation abilities (F_1,1210_ = 3.23, p = .020, η_p_^²^ = .008; Fig 6b).

**Fig 6.**
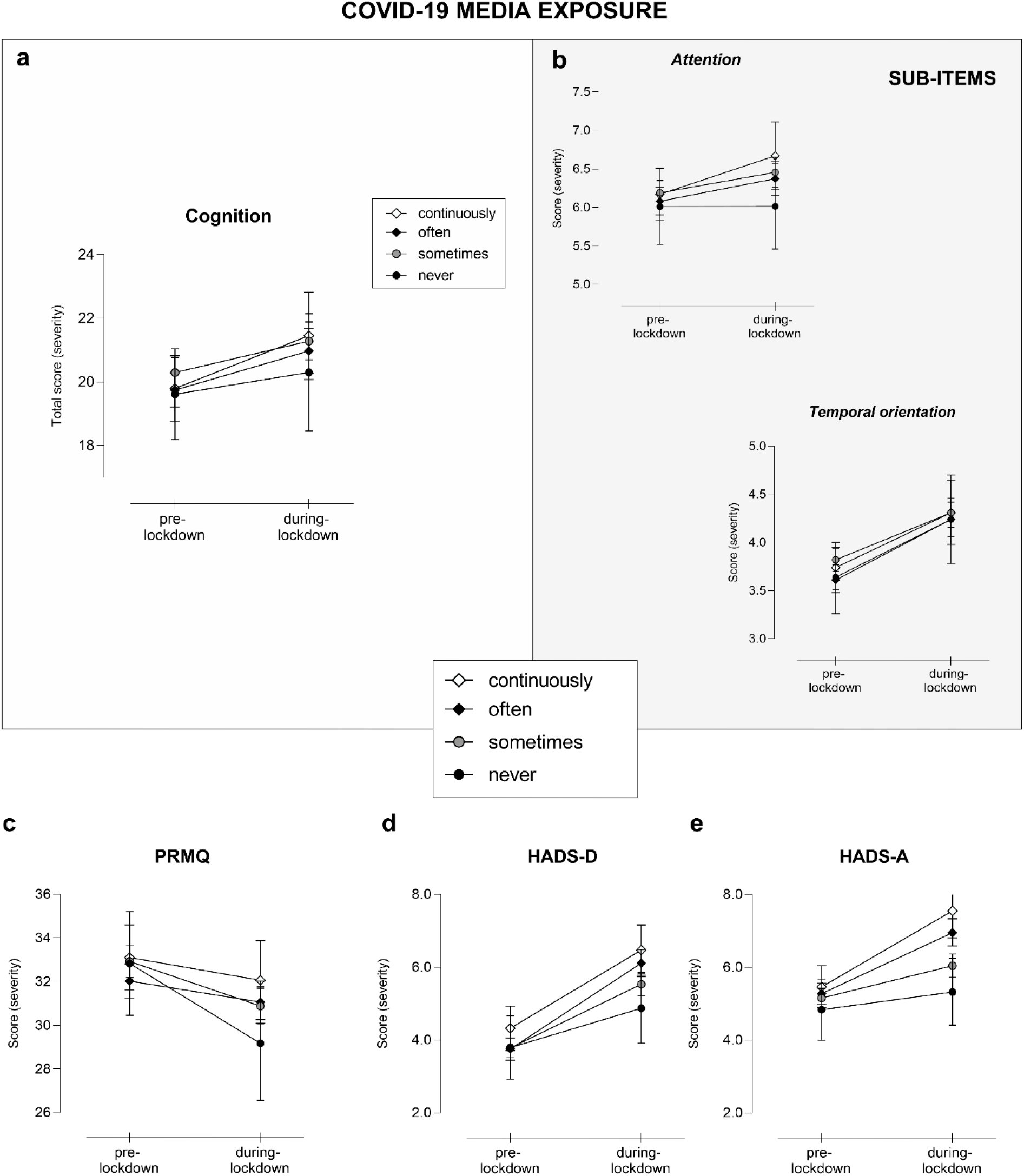
Lockdown effect according to COVID-19-media exposure. Changes in a) subjective cognitive functioning total score and b) its subitems, c) PRMQ, d) HADS-D and e) HADS-A as a function of Lockdown and Working condition. Only significant interactions Lockdown x COVID-19-media exposure are displayed, following RM-ANOVA analysis, entering age as covariate. Higher total score indicates higher severity in terms of cognitive and mental health symptoms. Error bars represent confidence intervals. PRMQ, Prospective and Retrospective Memory Questionnaire; HADS-D, Hospital Anxiety and Depression Scale for depression; HADS-A, HADS for anxiety.

Concerning the perceived improvement in memory domain, a statistically significant interaction Lockdown × Media exposure effect was found (F_1,1210_ = 4.60, p = .003, η_p_^²^ = .011; Fig 6c). More specifically, post-hoc analysis highlighted a subjective improvement during the lockdown as compared to normal times in the occasional media-seekers (never/sometimes) (p < .001), while no differences were perceived by the frequent media-seekers (often/ continuously) (p > .050).

About mental health, we noted a significant Lockdown × Media exposure interaction in HADS-D (F_1,1210_ = 5.63, p < .001, η_p_^²^ = .014; Fig 6d) and HADS-A (F_1,1210_ = 9.74, p < .001, η_p_^²^ = .024; Fig 6e), in terms of a higher level of depression and anxiety. Since post-hoc results showed mood and anxiety levels significantly changed in all the three groups (sometimes/often/continuously) (p < .001), we also conducted the planned comparisons to better investigate this interaction. This analysis revealed mood and anxiety worsening were greater in frequent media-seekers compared to occasional media-seekers (p < .050). Whereas changes in the other psychological behaviors were equally affected in all the working conditions.

## DISCUSSION

Did COVID-19-lockdown restrictions impact on perceived cognitive functioning as well as mental health? Current worldwide evidence underlines the exposure to this unprecedented stressful condition is increasing the prevalence of mental health disorders, as depression and anxiety. Our findings corroborate this evidence showing a worsening on mental health disorders. Notably, for the first time our study demonstrates that COVID-19-lockdown has a substantial impact on self-reported cognitive functioning. The results of the study allowed distinct vulnerability factors to be identified that are associated with higher risk of experiencing cognitive worsening and mental health disorders during the pandemic lockdown.

### Perceived cognitive functioning changes

Overall, our study showed the Italian lockdown restrictions had a detrimental effect on subjective cognitive functioning in its general population. Specifically, we identified the following risk factors of cognitive worsening related to COVID-19 lockdown – female gender, younger age, home confinement (due to teleworking or unemployment conditions) and repeated COVID-19-media consumption. In this regard, the subjective complaints were mostly perceived in everyday task involving attention, temporal orientation and executive functions; while no changes in language abilities were reported.

Conversely, about memory abilities, our findings revealed that working during the lockdown as well as not-frequently seeking for COVID-19 related information, were associated to a perceived improvement in memory domain (on both its prospective and retrospective components) compared to pre-lockdown times. Although, at first, this result can appear paradoxical, it is theoretically aligned with the established phenomenon of the ‘age-prospective memory paradox’ [26], wherein context changes were identified as a key factor in understanding the memory paradox. Here as well, the massive changes of the surrounding context and environment under COVID-19 lockdown play an important role in explaining our result. Changes of daily life routine under restrictions was characterized by a less frenetic rhythm and a reduction of possibilities, possibly minimizing the chances of experiencing memory failures and thus, leading to a subjective improvement of memory.

In this regard, a recent position paper emphasized the urgent need of collecting high-quality data on mental health, brain function, cognition of COVID-19 lockdown effects across the whole population [1], to identify vulnerable groups and possibly mitigate the psychological consequences under pandemic [2]. Our study extends previous knowledge and underlines the importance of considering these vulnerability factors to implement specific interventions by showing the presence of groups of individuals more likely to experience subjective cognitive changes.

Overall, the risk factors that we identified for subjective cognitive changes converge with those predicting psychological disturbances during COVID-19 pandemic [6–8,11,15] as well as previous isolation/ quarantine conditions [2,27].

Interestingly, being female was strongly associated with a higher level of cognitive complaints. Despite possible biological vulnerability factors for mental health issues and different stress responses related to the gender [28,29], in this unique context, a possible explanation could be that women (especially mothers) could get overloaded with family, household and/or working activities under restrictions [3]. This resulted in more difficulties to manage planned daytime activities due to the increased interaction between work and home lives, which in turn leads to further increases of stress level, lower self-efficacy and consequently poorer subjective cognitive functioning [30].

Furthermore, we identified being younger, home confinement (due to teleworking or unemployment conditions) and repeated COVID-19-media consumption as other vulnerability factors related to perceived cognitive worsening. Of note, young adults have already been identified by previous studies as more vulnerable to mental health disorders under COVID-19 pandemic [10,11]. Differently from older generation, young adults might experience additional stress due to future uncertainty (i.e., delays in academic career, job insecurity) and sense of loneliness, indeed profound changes occurred in their social and lifestyle habits leading to longer time spent on social media [31]. By contrast older adults, particularly elderly and retired people, under restrictions perceived a less pronounced change of their routine and social life, which is generally characterized by less social interactions and more time alone during daily activities, as compared to younger generations [32].

In line with these observations, occupational condition was identified as another vulnerability factor. Cognitive worsening was experienced by individuals being unemployed or in teleworking/studying, but not by regular workers. This finding suggests that the ‘confined at home’ working conditions were identified as more vulnerable groups, who possibly were more exposed to troubles, financial distress, fear of losing the job, changes in working habits and reductions on work-related social interactions.

Importantly, here for the first time we found repeated COVID-19-media exposure has an impact on subjective cognition – longer media exposure was related to higher level of cognitive complaints. Indeed, frequent media exposure to infectious diseases information can exacerbate stress responses and anxiety, amplify worries, ruminations and consequently impair functioning [2,33]. Our findings extend the current knowledge, by showing repeated pandemic-related media consumption can have detrimental effect on subjective cognition, along with higher levels of depression and anxiety (as shown in the next paragraph). Cognitive complaints might (also) be the result of high levels of distress and reduced wellbeing; indeed, over-exposure to anxiety-provoking media can lead to negative over-thinking, ruminations, continuous self-check-ups of COVID-19 related symptoms, which in turns keeps the mind busy with thoughts and worries leading to a worsening effect on cognitive abilities, negative perception of social dynamic and daily life functioning. This result emphasized the importance of understanding the effect of repeated COVID-19 related media consumption, to optimize health messaging around COVID-19 and develop specific guidelines to regulate media exposure in the context of this pandemic.

As previously mentioned, sub-items analysis revealed cognitive complaints mostly involved attentive abilities, executive functions and temporal orientation. Although in the literature there is no previous evidence about quarantine/lockdown effects on cognitive outcomes, few studies linked social isolation with adverse health consequences including impaired executive functions and faster cognitive decline along with depression, poor sleep, impaired immunity and poor cardiovascular function [34]. Concerning temporal orientation, our data corroborate previous findings, showing the lockdown induced a significant difficulty in keeping track of time, causing confusion about the exact day, week, month and time [35].

By contrast, regarding memory abilities, we found a paradoxical effect with people experiencing an overall improvement under COVID-19 restrictions. Namely, factors such as having a job (outside home or teleworking) and short exposure to COVID-19 outbreak information were predictive of more effective memory abilities, characterized by reduced memory slips in daily life activities under restrictions as compared to normal times. As mentioned in the previous paragraph, a possible explanation can be identified with the important context changes related to COVID-19 restrictions, where regular workers maintained their job but living with a less frenetic rhythm and a reduction of possibilities, as well as less chances of facing memory slips, leading in turns to a subjective improvement of their memory functioning. Conversely, individuals confined at home and without a job, did not perceive this improvement, possibly due to their more stressful condition with fear of losing their job, financial distress and future uncertainty. In this regard, we found subjective memory improvements were moderately associated with lower depressive and anxiety symptoms.

Finally, we used self-reported measures to assess cognitive functioning due to the nature of the study. Interestingly, subjective measures of cognitive functioning, but not objective, have been extensively recognized as significant predictors of everyday-task functioning (as assessed by ADL and IADL scales) [17], emphasizing their strong validity and potential clinical value [36]. Further, a strength of our study design is that we included a 2-time points assessment (pre-vs. during-lockdown), allowing a direct comparison of the self-reported measures within-subject and hence, minimizing possible variability issues.

### Mental health changes

In relation to the second aim, an important expected finding of our study, was the adverse impact on mental health and other psychological behaviors, associated with the COVID-19-prolonged lockdown. Specifically, higher levels of anxiety and depressive disorders were observed, as well as significant changes in sleeping pattern, appetite, interest in sex and health-related anxiety.

Indeed, during the last weeks of Italian lockdown restrictions, mild/severe levels of depression increased up to the 32.30% compared to 15.39% before the lockdown, which are in line with the results of other Italian [8] as well as Chinese studies [10], suggesting a sharp increase of depression level in the general population. Interestingly, this higher prevalence was driven mostly by a significant increment of moderate/severe cases, which raised up to 13.71% under restrictions as compared to the 4.35% during normal conditions.

Regarding anxiety, under lockdown the prevalence of mild/severe levels of anxiety increased up to the 35.72% compared to the 21.40% before the lockdown. These results corroborate previous studies in the Italian [37] as well as Chinese population [11,38]. Again, we noted that the higher prevalence of anxiety disturbances under restriction was mainly driven by a significant increment of moderate/severe cases, which raised up to 16.86% under restrictions as compared to the 6.69% before the lockdown.

Our results emphasized that under restrictions, the increase of depressive and anxiety disorders was not only in terms of prevalence, but also clinical severity. In this context of COVID-19 pandemic, monitoring severity seems particularly important, as there is the urgent need to early detect milder clinical cases and implement efficacious intervention, before they evolve to more complex and stable clinical profiles [15].

In addition, the lockdown had a negative impact on sleep disorders (with people experiencing insomnia or increased sleepiness), which has been extensively reported in the literature [3,14,35,37]. Other relevant psychological issues under restrictions included: increased appetite, reduced libido and higher level of anxiety for health, corroborating previous evidence [39–41]. Among these issues, eating more seems to be a frequent and natural reaction under certain stressful situations [42]; however, it is worth mentioning that combined with home confinement and reduced motor activity can lead to other health risks.

Taken together, these findings confirmed COVID-19 pandemic had a significant impact on mental health issues; and to better understand this phenomenon, we further explored its interaction with relevant factors which could modulate changes in mental health disorders.

About depression, we identified distinct risk factors: namely, female gender, younger age (particularly age ranges under 45 years), unemployment status, being resident in Northern Italy and repeated COVID-19-media consumption, which resulted as influential variables for the development of depressive symptoms.

Similarly, for anxiety: being female, unemployed and a frequent COVID-19-media seeker, were identified as relevant factors in predicting anxiety disorders under lockdown. As stressed by our results and confirmed by other European studies, women have been depicted as more vulnerable to depression and anxiety disorders than men under COVID-19 lockdown [6,8]. By contrast in China an opposite scenario was reported [5,11]. Our study highlighted an important gender difference associated with lockdown restrictions, showing the probability of developing depression was 1.6-fold and for anxiety 2.2-fold greater in women than men. In the context of no differences for depression between genders before the lockdown and only slight for anxiety. Differently from men, during the lockdown, women experienced a change also in terms of appetite, which increased for the majority of the sample, as well as reported higher levels of health-related anxiety than normal times (pre-lockdown). However, we did not find any difference between genders in changes in libido and differently from previous study in sleep disturbances [7].

Despite the already established gender gap showing higher prevalence of mental health disorders in women [28], overall our data highlight the severe impact of lockdown on psychological well-being, with women being the most vulnerable.

Young adults resulted as at higher risk of experiencing depressive and anxiety disorders, as well as other psychological issues than older generations (>45 years). Namely, youth were more likely to experience sleeping disorders (i.e., the 26-45 aged group), increased eating behaviors, changes in sexual desire and increased hypochondria. At first, this could seem a counterintuitive result as COVID-19 fatality rate is higher among elderly [43], however our findings are in line with previous evidence, which identified young adults as a vulnerable population for psychological issues associated to COVID-19 pandemic lockdown [7,10,11,44]. Indeed, younger people are exposed to: future uncertainty related to prolonged academia closure and precarious employments, substantial changes in social and lifestyle with more time spent on social media – all aspects, which can easily trigger stress, leading to mental health issues [5,31,45].

As expected, being unemployed was associated with higher level of depression and anxiety under restrictions, in this regard in our sample, we found an overall reduction of about 1.5 daily working hours [6.75(3.37) vs. 5.03 (3.78), F_1,1214_=329.92, p < .001, η_p_^²^ = .214].

However, differently from previous evidence we did not find regular workers, which includes healthcare professionals, to be at higher risk for mental illness [7,13,15,38]. We believe this is due to the low representativeness of front-line healthcare workers in our sample (only 7.9%), with the majority of regular workers not experiencing troubles or financial insecurity as compared to the unemployed group [8,46].

Another risk factor has been identified in the Italian region of residence, namely living in Northern Italy under COVID-19 pandemic was associated with increased mental health disturbances: higher level of depression and hypochondria (i.e., anxiety for health). Indeed, Northern Italy was considered as the core region of the emergency – with the highest number of infections and deaths as well as where the outbreak started. Our results are consistent with previous pandemics evidence (e.g., SARS), showing residents of the most infected regions had a higher risk of developing mental health disorders, particularly depression [47]. Further, about the increased level of anxiety for health, a possible explanation is people living in regions with high infection-prevalence can perceive themselves as at higher risk to contract the disorder [14].

Importantly, among risk factors for depression and anxiety, we identified repeated COVID-19-media exposure. Our results partially agree with a recent study from China, showing elevated levels of anxiety, but not depression, in association to the increased time spent on following COVID-19 news [11]. Of note, anxiety and uncertainty can lead to further media consumption and additional distress, generating a vicious cycle [48]. In addition, spending increased time on internet during the lockdown has been linked with depression disorders [7]. In this regard, since overexposure to COVID-19-media amplifies distress, leading to mental illness as well as cognitive complaints, future research and guidelines are needed to promote wellbeing by indicating the optimal patterns of media consumption under a pandemic [45].

### Limitations and strengths

There are a few shortcomings and limitations in the current work that have to be considered. First, the survey sampling was based on the snowball method, involving an online invitation, but leaving unexplored the population who did not use network devices. Furthermore, considering the online distribution, no data about people who refused to participate were collected and no refusal rate was registered. However, during home confinement this was the only feasible sampling method. Moreover, to collect a heterogeneous sample we encouraged participants to invite new respondents by involving elderly and people with a poor access to internet. Indeed, our sample seems to have an adequate representation of the Italian general population in terms of age (range: 18-88 years), geographical distribution across Italy and educational level, but less accurate for gender, as ∼70% of the sample was female. Second, subjective complaints and mental health outcomes are based on self-reported measures rather than clinical diagnoses, although the majority of the selected scales were validated [18,24] or derived from standardized tools [19]. In particular, depression and anxiety prevalence were based on HADS, which has excellent psychometric properties and is extensively used in the general clinical practice [22]. Finally, although our study has not a longitudinal design, it comprised a pre-lockdown assessment, which was used as a reference to conduct pre-post analyses with the data collected during the lockdown. Therefore, differently from previous evidence on mental health [15], we could infer the observed changes were possibly due the COVID-19-lockdown effect.

## CONCLUSIONS

This is the first known study showing lockdown can have a severe impact on subjective cognitive functioning, along with mental health disorders. Importantly, we have shown under restrictions people can experience subjective cognitive complaints in attentive, executive, temporal orientation abilities, while paradoxically perceiving an improvement in memory domain, with a reduction in memory slips in daily functioning. Finally, we found higher severity and prevalence of depression, anxiety disorders and other psychological issues involving sleep, appetite, libido and hypochondria.

Hence, considering home confinement can be reimposed during COVID-19 pandemic, knowing the associated cognitive and psychological effects is crucial to provide effective and supportive psychological interventions, particularly to vulnerable populations, which as outlined in our results can be characterized by the following influential risk factors: being female, younger age (<45 years), unemployment status, being resident in high infection-prevalence areas and a repeated COVID-19-media exposure.

To conclude, we believe future researches is needed to define the long-term consequences of an epidemic lockdown on subjective cognition and mental health disorders, as well as to define specific guidelines especially about COVID-19-media consumption in order to minimize the psychological impact associated to this pandemic.

## Data Availability

All relevant data are within the manuscript. Data cannot be shared publicly due to ethical restrictions, as we have no permission from the participants of our survey to share the de-identified dataset with the general public. Readers can contact to request the data.

## SUPPLEMENTARY MATERIAL

**S1 Fig.**
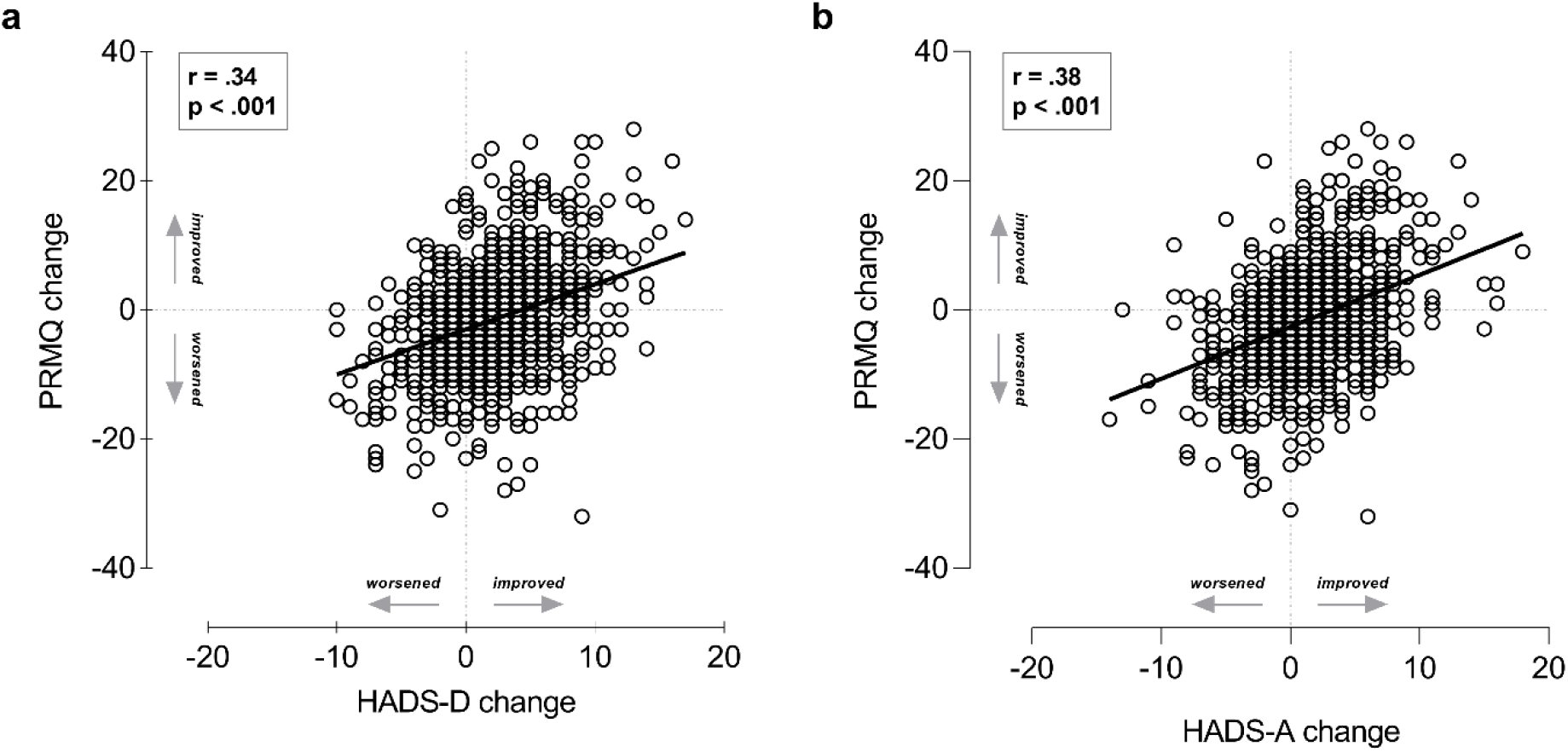
Changes in Prospective and Retrospective Memory Questionnaire (PRMQ) score as a function of changes in: a) depression (HADS-D) and b) anxiety (HADS-A). Changes scores were obtained by subtracting before lockdown score from during lockdown). HADS-D, Hospital Anxiety and Depression Scale for depression; HADS-A, HADS for anxiety.

**S1 Table.**
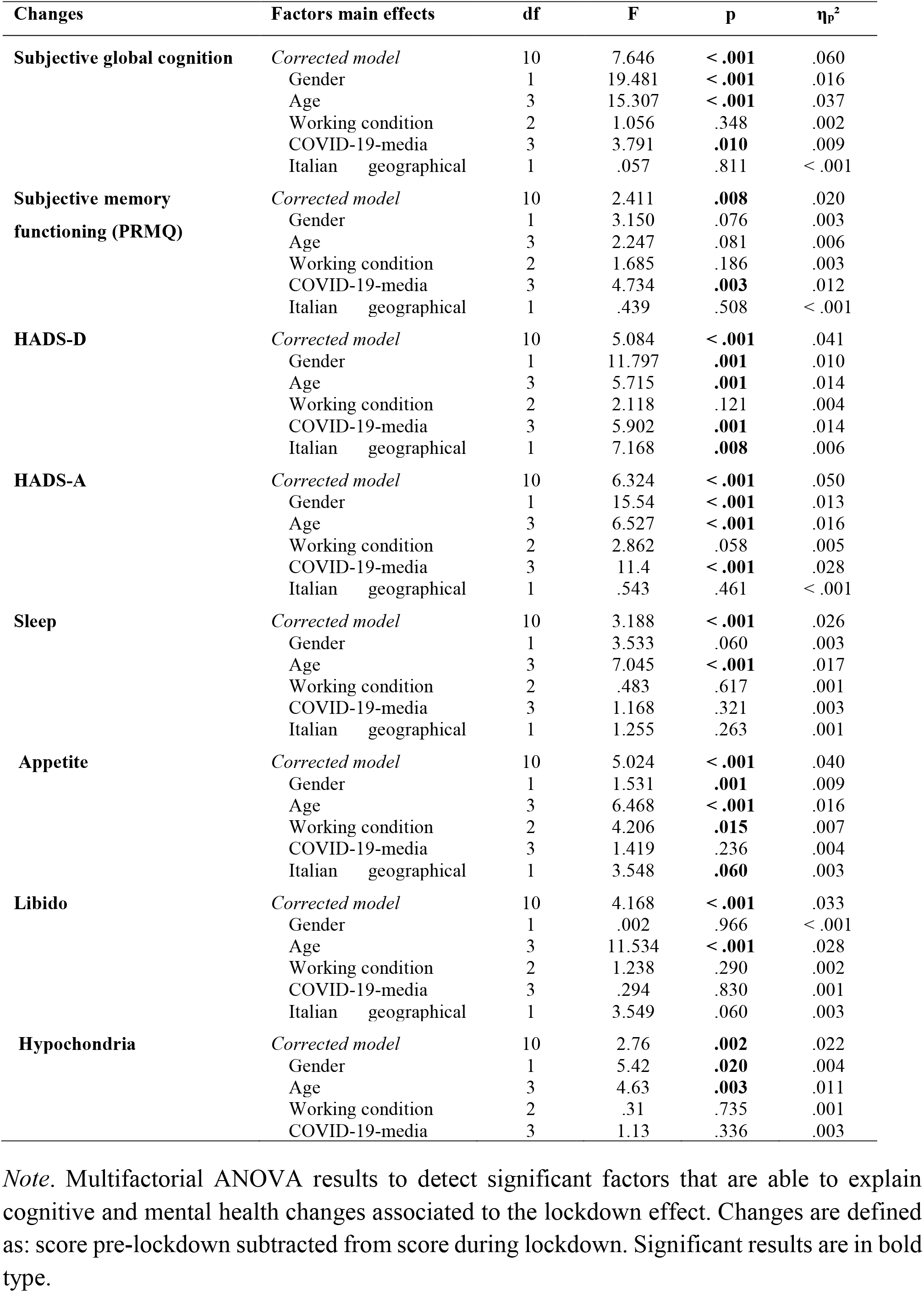
Multi-factorial models.

## S1 APPENDIX

Questionnaire to assess global cognitive functioning and items categorization according to the involved cognitive functions

INSTRUCTIONS: Please answer all questions as accurately as possible, indicating how often the following situations occurred to you (very often, quite often, sometimes, rarely, never).

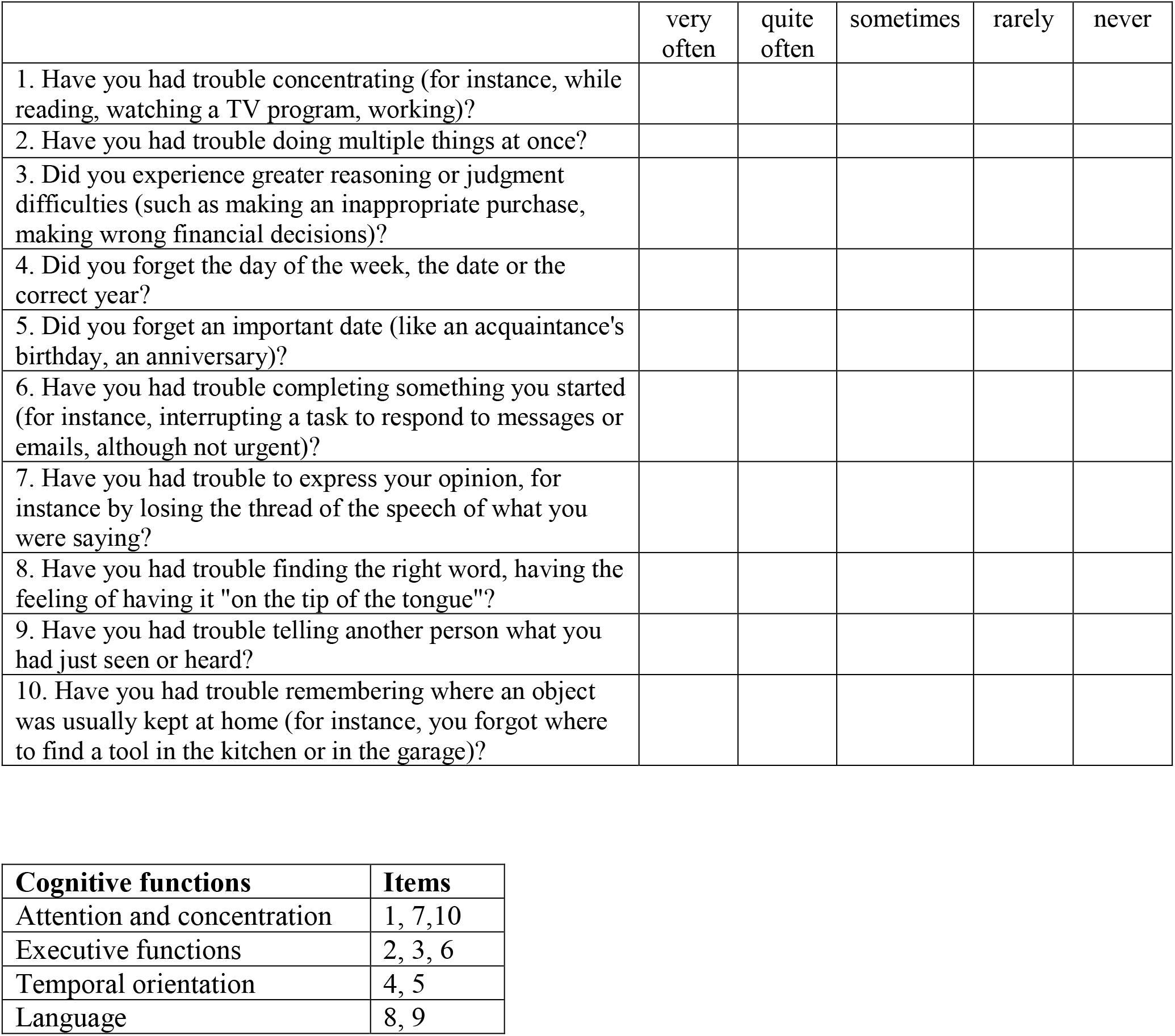

## REFERENCES

1. Holmes EA, O’Connor RC, Perry VH, Tracey I, Wessely S, Arseneault L, et al. Multidisciplinary research priorities for the COVID-19 pandemic: a call for action for mental health science. Lancet Psychiatry. 2020;7: 547–560.

2. Brooks SK, Webster RK, Smith LE, Woodland L, Wessely S, Greenberg N, et al. The psychological impact of quarantine and how to reduce it: rapid review of the evidence. The Lancet. 2020;395: 912–920.

3. Altena E, Baglioni C, Espie CA, Ellis J, Gavriloff D, Holzinger B, et al. Dealing with sleep problems during home confinement due to the COVID-19 outbreak: Practical recommendations from a task force of the European CBT-I Academy. J Sleep Res. 2020;29:p e13052.

4. Ho CS, Chee CY, Ho RC. Mental Health Strategies to Combat the Psychological Impact of Coronavirus Disease 2019 (COVID-19) Beyond Paranoia and Panic. 2020;49: 6.

5. Wang C, Pan R, Wan X, Tan Y, Xu L, Ho CS, et al. Immediate Psychological Responses and Associated Factors during the Initial Stage of the 2019 Coronavirus Disease (COVID-19) Epidemic among the General Population in China. Int J Environ Res Public Health. 2020;17: 1729.

6. González-Sanguino C, Ausín B, Castellanos MÁ, Saiz J, López-Gómez A, Ugidos C, et al. Mental health consequences during the initial stage of the 2020 Coronavirus pandemic (COVID-19) in Spain. Brain Behav Immun. 2020;87: 172–176.

7. Gualano MR, Lo Moro G, Voglino G, Bert F, Siliquini R. Effects of Covid-19 Lockdown on Mental Health and Sleep Disturbances in Italy. Int J Environ Res Public Health. 2020;17:4779.

8. Mazza C, Ricci E, Biondi S, Colasanti M, Ferracuti S, Napoli C, et al. A Nationwide Survey of Psychological Distress among Italian People during the COVID-19 Pandemic: Immediate Psychological Responses and Associated Factors. Int J Environ Res Public Health. 2020;17: 3165.

9. Tull MT, Edmonds KA, Scamaldo KM, Richmond JR, Rose JP, Gratz KL. Psychological Outcomes Associated with Stay-at-Home Orders and the Perceived Impact of COVID-19 on Daily Life. Psychiatry Res. 2020;289: 113098.

10. Ahmed MZ, Ahmed O, Aibao Z, Hanbin S, Siyu L, Ahmad A. Epidemic of COVID-19 in China and associated Psychological Problems. Asian J Psychiatry. 2020;51: 102092.

11. Huang Y, Zhao N. Generalized anxiety disorder, depressive symptoms and sleep quality during COVID-19 outbreak in China: a web-based cross-sectional survey. Psychiatry Res. 2020;288: 112954.

12. Qiu J, Shen B, Zhao M, Wang Z, Xie B, Xu Y. A nationwide survey of psychological distress among Chinese people in the COVID-19 epidemic: implications and policy recommendations. Gen Psychiatry. 2020;33: e100213.

13. Zhang C, Yang L, Liu S, Ma S, Wang Y, Cai Z, et al. Survey of Insomnia and Related Social Psychological Factors Among Medical Staff Involved in the 2019 Novel Coronavirus Disease Outbreak. Front Psychiatry. 2020;11: 306.

14. Casagrande M, Favieri F, Tambelli R, Forte G. The enemy who sealed the world: Effects quarantine due to the COVID-19 on sleep quality, anxiety, and psychological distress in the Italian population. Sleep Med. 2020;75: 12–20.

15. Pappa S, Ntella V, Giannakas T, Giannakoulis VG, Papoutsi E, Katsaounou P. Prevalence of depression, anxiety, and insomnia among healthcare workers during the COVID-19 pandemic: A systematic review and meta-analysis. Brain Behav Immun. 2020; 88, 901–907.

16. Reisberg B, Shulman MB, Torossian C, Leng L, Zhu W. Outcome over seven years of healthy adults with and without subjective cognitive impairment. Alzheimers Dement. 2010;6: 11–24.

17. Song M-K, Ward SE, Bair E, Weiner LJ, Bridgman JC, Hladik GA, et al. Patient-reported cognitive functioning and daily functioning in chronic dialysis patients: Cognitive and daily functioning in dialysis patients. Hemodial Int. 2015;19: 90–99.

18. Crawford J, Smith G, Maylor E, Sala SD, Logie R. The Prospective and Retrospective Memory Questionnaire (PRMQ): Normative data and latent structure in a large non-clinical sample. Memory. 2003;11: 261–275.

19. Sullivan MJ, Edgley K, Dehoux E. A survey of multiple sclerosis: I. Perceived cognitive problems and compensatory strategy use. Can J Rehabil. 1990;4: 99–105.

20. Damin AE, Nitrini R, Brucki SMD. Cognitive Change Questionnaire as a method for cognitive impairment screening. Dement Neuropsychol. 2015;9: 237–244.

21. Zigmond, A.S., Snaith, R.P., 1983. The Hospital Anxiety and Depression Scale. Acta Psychiatr. Scand. 67, 361–370.

22. Olssøn I, Mykletun A, Dahl AA. The hospital anxiety and depression rating scale: A cross-sectional study of psychometrics and case finding abilities in general practice. BMC Psychiatry. 2005;5: 46.

23. Costantini M, Musso M, Viterbori P, Bonci F, Del Mastro L, Garrone O, et al. Detecting psychological distress in cancer patients: validity of the Italian version of the Hospital Anxiety and Depression Scale. Support Care Cancer. 1999;7: 121–127.

24. Beck AT, Steer RA, Brown GK. Beck depression inventory (BDI-II). Pearson; 1996.

25. Hamilton M. The Hamilton rating scale for depression. Assessment of depression. Springer; 1986. pp. 143–152.

26. Schnitzspahn KM, Ihle A, Henry JD, Rendell PG, Kliegel M. The age-prospective memory-paradox: an exploration of possible mechanisms. Int Psychogeriatr. 2011;23: 583–592.

27. Taylor MR, Agho KE, Stevens GJ, Raphael B. Factors influencing psychological distress during a disease epidemic: Data from Australia’s first outbreak of equine influenza. BMC Public Health. 2008;8: 347.

28. Albert PR. Why is depression more prevalent in women? J Psychiatry Neurosci JPN. 2015;40: 219–221.

29. Young E, Korszun A. Sex, trauma, stress hormones and depression. Mol Psychiatry. 2010;15: 23–28.

30. Albert MS, Jones K, Savage CR, Berkman L, Seeman T, Blazer D, et al. Predictors of cognitive change in older persons: MacArthur studies of successful aging. Psychol Aging. 1995;10: 578–589.

31. Ozamiz-Etxebarria N, Idoiaga Mondragon N, Dosil Santamaría M, Picaza Gorrotxategi M. Psychological Symptoms During the Two Stages of Lockdown in Response to the COVID-19 Outbreak: An Investigation in a Sample of Citizens in Northern Spain. Front Psychol. 2020;11.

32. Marcum CS. Age Differences in Daily Social Activities. Res Aging. 2013;35: 612–640.

33. Thompson RR, Garfin DR, Holman EA, Silver RC. Distress, Worry, and Functioning Following a Global Health Crisis: A National Study of Americans’ Responses to Ebola. Clin Psychol Sci. 2017;5: 513–521.

34. Hawkley LC, Capitanio JP. Perceived social isolation, evolutionary fitness and health outcomes: a lifespan approach. Philos Trans R Soc B Biol Sci. 2015;370: 20140114.

35. Cellini N, Canale N, Mioni G, Costa S. Changes in sleep pattern, sense of time and digital media use during COVID-19 lockdown in Italy. J Sleep Res. 2020;29: e13074

36. Chelune GJ, Heaton RK, Lehman RA. Neuropsychological and personality correlates of patients’ complaints of disability. Advances in clinical neuropsychology. New York: Plenum Press; 1986. pp. 95–126.

37. Marelli S, Castelnuovo A, Somma A, Castronovo V, Mombelli S, Bottoni D, et al. Impact of COVID-19 lockdown on sleep quality in university students and administration staff. J Neurol. 2020. doi:10.1007/s00415-020-10056-6

38. Huang Y, Zhao N. Chinese mental health burden during the COVID-19 pandemic. Asian J Psychiatry. 2020;51: 102052.

39. Di Renzo L, Gualtieri P, Pivari F, Soldati L, Attinà A, Cinelli G, et al. Eating habits and lifestyle changes during COVID-19 lockdown: an Italian survey. J Transl Med. 2020;18: 229.

40. Di Renzo L, Gualtieri P, Cinelli G, Bigioni G, Soldati L, Attinà A, et al. Psychological Aspects and Eating Habits during COVID-19 Home Confinement: Results of EHLC-COVID-19 Italian Online Survey. Nutrients. 2020;12: 2152.

41. Ko N-Y, Lu W-H, Chen Y-L, Li D-J, Chang Y-P, Wu C-F, et al. Changes in Sex Life among People in Taiwan during the COVID-19 Pandemic: The Roles of Risk Perception, General Anxiety, and Demographic Characteristics. Int J Environ Res Public Health. 2020;17: 5822.

42. Razzoli M, Pearson C, Crow S, Bartolomucci A. Stress, overeating, and obesity: Insights from human studies and preclinical models. Neurosci Biobehav Rev. 2017;76: 154–162.

43. Liu K, Chen Y, Lin R, Han K. Clinical features of COVID-19 in elderly patients: A comparison with young and middle-aged patients. J Infect. 2020;80: e14–e18.

44. Yuan S, Liao Z, Huang H, Jiang B, Zhang X, Wang Y, et al. Comparison of the Indicators of Psychological Stress in the Population of Hubei Province and Non-Endemic Provinces in China During Two Weeks During the Coronavirus Disease 2019 (COVID-19) Outbreak in February 2020. Med Sci Monit Int Med J Exp Clin Res. 2020;26: e923767-1-e923767-10.

45. Garfin DR, Silver RC, Holman EA. The novel coronavirus (COVID-2019) outbreak: Amplification of public health consequences by media exposure. Health Psychol. 2020;39: 355–357.

46. Lei L, Huang X, Zhang S, Yang J, Yang L, Xu M. Comparison of Prevalence and Associated Factors of Anxiety and Depression Among People Affected by versus People Unaffected by Quarantine During the COVID-19 Epidemic in Southwestern China. Med Sci Monit. 2020;26.

47. Lee TMC, Chi I, Chung LWM, Chou K-L. Ageing and psychological response during the post-SARS period. Aging Ment Health. 2006;10: 303–311.

48. Thompson RR, Jones NM, Holman EA, Silver RC. Media exposure to mass violence events can fuel a cycle of distress. Sci Adv. 2019;5: eaav3502.

